# Study protocol Effects of Philips Visual Patient Avatar on vital sign deviations and audible alarm burden in perioperative care: a dual-centre, quasi-experimental pre-post big-data study protocol (NewYork-Presbyterian/Weill Cornell and University Hospital Zurich)

**DOI:** 10.64898/2026.05.18.26353454

**Authors:** Silis Y. Jiang, Tadzio R. Roche, Krzysztof Cybulski, Gaspar Dugac, Lukas Meier, Virginia E. Tangel, Max Ebensperger, Andreas Maskos, Michael Tucci, Christoph B. Nöthiger, Markus Kalisch, Zachary A. Turnbull, David W. Tscholl

## Abstract

**Introduction:** Perioperative patient monitoring requires clinicians to integrate multiple physiological data streams under time pressure and frequent interruptions. Conventional monitors predominantly present vital signs as separate numerical values and waveforms, which must be sequentially interpreted and mentally integrated, imposing substantial cognitive demands. Audible alarms are intended to enhance safety but contribute to alarm fatigue and increased workload. Time spent outside predefined safe ranges for key physiological variables and excessive alarm burden are associated with adverse outcomes, motivating approaches that support earlier detection and improved situation awareness without increasing cognitive load.

The Philips Visual Patient Avatar is an avatar-based visualisation technology displayed on the patient monitor that supports clinicians’ situation awareness by integrating multiple vital signs and sensor states into a single animated virtual patient, while retaining conventional numerical displays.

Although laboratory, simulation and qualitative studies suggest benefits of avatar-based monitoring, its impact on objective monitoring outcomes has not been systematically quantified.

**Methods and analysis:** This investigator-initiated, dual-centre, quasi-experimental pre–post study evaluates the clinical introduction of VPA using routinely collected perioperative monitoring data. The study is conducted at NewYork-Presbyterian Hospital / Weill Cornell Medical Center (New York, USA) and University Hospital Zurich (Zurich, Switzerland).

At each centre, data are collected across three phases: pre-implementation, a sink-in (adaptation) phase during staged installation and training, and post-implementation. The unit of analysis is the individual anaesthesia case. Outcomes are analysed retrospectively.

The primary outcome is vital-sign deviation time, normalised to 60 minutes of case time, defined as cumulative time outside predefined upper or lower thresholds for each vital sign. Secondary outcomes include a time-based area-under-the-curve metric capturing magnitude and duration of deviations, as well as audible alarm burden (alarm count, alarm time and alarm reaction time). Multivariable regression models will estimate associations between VPA implementation phase and outcomes while adjusting for relevant patient- and procedure-related covariates. Analyses will be performed separately for each centre, with sensitivity analyses as appropriate.

**Ethics and dissemination:** The study was approved by the Institutional Review Board of Weill Cornell Medicine. For Zurich, a declaration of non-jurisdiction was issued for the analysis of anonymised routinely collected data. At the time of protocol submission, data collection is complete at University Hospital Zurich and ongoing at NewYork-Presbyterian Hospital / Weill Cornell Medical Center; no analyses have been performed. Findings will be disseminated through peer-reviewed publications and scientific conferences.

**Trial registration:** Not applicable (observational analysis of routinely collected data).

**Strengths and limitations of this study:**

- This dual-centre study leverages large volumes of routinely collected perioperative monitoring data from two tertiary academic hospitals on different continents, enhancing external validity across diverse clinical settings.
- The quasi-experimental pre–post design enables evaluation of the Visual Patient Avatar under routine clinical conditions but does not permit causal inference.
- A dedicated sink-in phase was included to mitigate learning and adaptation effects; however, residual confounding by time and unmeasured factors cannot be fully excluded.
- Differences in alarm configurations and visualisation thresholds between centres reflect local standards of care and limit direct standardisation, while allowing assessment of the intervention across heterogeneous real-world alarm environments.
- Outcomes focus on process-level monitoring and alarm metrics rather than hard patient outcomes; however, deviations in vital signs and alarm burden are supported by prior evidence as clinically meaningful indicators of patient safety. If improvements are observed, this study will provide the basis for future investigations evaluating patient-centred clinical outcomes.

## Introduction

Continuous physiological patient monitoring is universally regarded as a cornerstone of safe anaesthesia and surgery, forming the backbone of perioperative patient safety(1–3). Perioperative patient monitoring requires clinicians to continuously integrate multiple physiological data streams (e.g., heart rate, blood pressure, oxygen saturation, and end-tidal CO2) under time pressure, frequent interruptions and competing tasks. Conventional patient monitors typically present vital signs as separate numerical values and waveforms, which clinicians must sequentially read and mentally integrate into an overall assessment of the patient’s physiological state. This process can impose substantial cognitive demands, particularly in dynamic, high-acuity clinical environments(4–12).

Audible alarms are intended to alert clinicians to potentially hazardous situations but are also a major contributor to alarm fatigue. High alarm rates and prolonged alarm sounding may distract clinicians, increase cognitive workload and reduce responsiveness to clinically relevant events(13–15). Beyond alarm burden, time spent outside predefined safe ranges for key physiological variables—such as intraoperative hypotension—has been associated with adverse outcomes in perioperative and critical care research(16–18). These findings motivate efforts to detect physiological deviations earlier and to support timely corrective actions without increasing cognitive load.

The Philips (Koninklijke Philips N.V., Amsterdam, The Netherlands) Visual Patient Avatar (VPA) is an avatar-based patient monitoring visualisation designed to support clinicians’ situation awareness(8, 11, 19–21). The system displays multiple vital signs and sensor states simultaneously on a single animated virtual patient, while conventional numerical values and waveforms remain available. Changes in physiological status are conveyed through intuitive visual cues, such as skin colour to represent oxygen saturation or pulsation dynamics to represent pulse rate and blood pressure.

This approach builds on principles of situation-awareness–oriented design and human visual perception by integrating multiple physiological parameters into a single, anatomically meaningful visual representation(22). By mapping vital signs and sensor states to intuitive visual features such as colour, shape, motion and spatial location, the system supports rapid perception, holistic comprehension and pattern recognition of the patient’s physiological state. This design reduces reliance on sequential numerical interpretation and active mental integration, and is intended to support clinicians’ situation awareness across varying levels of workload, attention and viewing distance.

### Evidence gap and rationale

Evidence from laboratory experiments, simulation studies and qualitative clinical investigations suggests that avatar-based patient monitoring can improve information transfer, increase diagnostic confidence and reduce perceived workload compared with conventional monitoring displays(9, 10, 12, 23–29). However, despite growing preclinical and simulation-based evidence, the real-world impact of VPA on objective monitoring outcomes in routine perioperative care has not yet been systematically quantified.

This study protocol describes a dual-centre, big-data evaluation conducted at NewYork-Presbyterian Hospital/Weill Cornell Medical Center (New York, USA) and University Hospital Zurich (Zurich, Switzerland). The study aims to assess whether the clinical introduction of VPA in routine perioperative practice is associated with (1) reduced time spent outside predefined vital-sign thresholds and (2) a reduced burden of audible alarms across diverse patient populations, clinical environments and care areas, including operating rooms, post-anaesthesia care units and procedural areas.

## Methods and analysis

### Study design

This study is an investigator-initiated, dual-centre, quasi-experimental pre–post study using routinely collected perioperative monitoring data to evaluate the real-world clinical impact of the VPA situation awareness system.

At each centre, the study follows a three-phase design consisting of (1) a pre-implementation phase, (2) a sink-in (adaptation) phase during staged installation and training, and (3) a post-implementation phase after completion of installation. This structure allows assessment of changes associated with VPA implementation under routine clinical conditions while accounting for learning effects and gradual adoption.

The unit of analysis is the individual anaesthesia case. By including large case volumes from two independent tertiary academic hospitals on different continents, the study aims to enhance the robustness, external validity and reproducibility of the findings across diverse perioperative settings.

Although outcome data are analysed retrospectively, the intervention and study phases were defined prospectively based on the implementation timeline at each centre.

### Rationale for the pre–post study design

A pre–post design was chosen as the most appropriate methodological approach to evaluate the effects of VPA in routine clinical practice. Randomisation at the patient or provider level was not feasible because the intervention is implemented at the level of the monitoring infrastructure and affects entire care areas simultaneously.

Leveraging routinely collected large-scale clinical data enables evaluation of the intervention without disrupting established workflows and allows assessment across a broad spectrum of patient populations, care areas and clinical contexts.

### Patient and public involvement

Patients and the public were not involved in the design, conduct, reporting or dissemination plans of this study.

### Study setting and participating centres

The study is conducted at two tertiary academic hospitals:

- NewYork-Presbyterian Hospital / Weill Cornell Medical Center (NYP/WCM), New York, USA, a large academic medical centre with over 4,000 beds and comprehensive perioperative services. NYP/WCM was the first centre in North America to clinically implement VPA.
- University Hospital Zurich (USZ), Zurich, Switzerland, a tertiary academic hospital with approximately 900 beds and the primary teaching hospital of the University of Zurich, situated within one of Europe’s leading academic and technological hubs alongside ETH Zurich. USZ was the first centre worldwide to implement VPA in routine clinical practice.

### Study phases and timeline

Phase timing differs between centres because regulatory approvals (CE, for Switzerland and FDA, for USA) occurred at different times. At each centre, the update of the first patient monitor marks the end of the pre-implementation phase and the beginning of the sink-in phase.

The sink-in phase represents a transition period during staged installation and training and allows for learning effects and gradual adoption. Its duration is defined by the local implementation schedule rather than by a fixed calendar interval.

Data are collected during three consecutive phases: pre-implementation, sink-in and post-implementation. Phase definitions, timing and expected case numbers for each centre are provided in **Table 1**. Overall, approximately 16,000 cases at NYP/WCM and 42,000 cases at USZ are expected to be included.

**Table 1:** Study phases, case numbers, and Visual Patient Avatar (VPA) deployment by centre.

|  | **NewYork-Presbyterian/Weill Cornell** | **University Hospital Zurich** |
| --- | --- | --- |
| Pre-phase timeframe | Sep 1, 2024 to Feb 14, 2025 (167 days) | Aug 31, 2022 to Jan 31, 2023 (154 days) |
| Sink-in phase timeframe | Mar 1, 2025 to Aug 31, 2025 (184 days) | Feb 18, 2023 to Aug 15, 2023 (179 days) |
| Post-phase timeframe | Sep 1, 2025 to Feb 28, 2026 (181 days) | Aug 16, 2023 to Feb 12, 2024 (181 days) |
| Approximate number of total cases | 16,000 | 42,000 |
| Total number of monitors with VPA in the operating rooms and post anaesthesia care units of the study centre | 49 | 126 |

At the time of submission of this protocol, data collection at the University Hospital Zurich (USZ) has been completed. Data collection at NewYork-Presbyterian Hospital / Weill Cornell Medical Center (NYP/WCM) is ongoing. No data analyses have been performed at either centre at the time of submission.

### Participating clinical care areas

The study includes perioperative care areas with differing workflows, professional groups and patient populations:

- Operating rooms (OR) at both centres
- Post-anaesthesia care units (PACU) at both centres
- Procedural areas at USZ only, including neuro-interventional radiology, interventional pulmonology and emergency trauma care

### Study participants (inclusion criteria)

- All paediatric and adult anaesthesia cases conducted in the participating care areas during the prespecified study phases with available perioperative monitoring data will be included.

Paediatric cases (≤18 years) will be analysed and reported separately in a dedicated publication. This separation is necessary because the participating centres apply different alarm thresholds and monitor configurations for paediatric populations and because physiological ranges and response patterns differ substantially from adults(30, 31). Pooling adult and paediatric data would therefore introduce clinically relevant heterogeneity.

### Intervention: VPA implementation

The intervention consists of the introduction of the VPA as a visual situation awareness system via a CE- and FDA-certified software update to Philips IntelliVue patient monitors used in the participating centres (i.e., MX450, MX550, and MX750).

The VPA is displayed alongside conventional numerical values and physiological waveforms in a split-screen view (**Figure 1** and **2**). The avatar visualisation is enabled by default, and no monitor configurations without VPA are available. Clinicians choose between several predefined screen layouts that differ in the arrangement and relative prominence of the avatar and conventional monitoring elements (**Supplementary Figure S1**). These layouts were selectable at the user level and could be changed at any time during routine clinical care.

**Figure 1:**
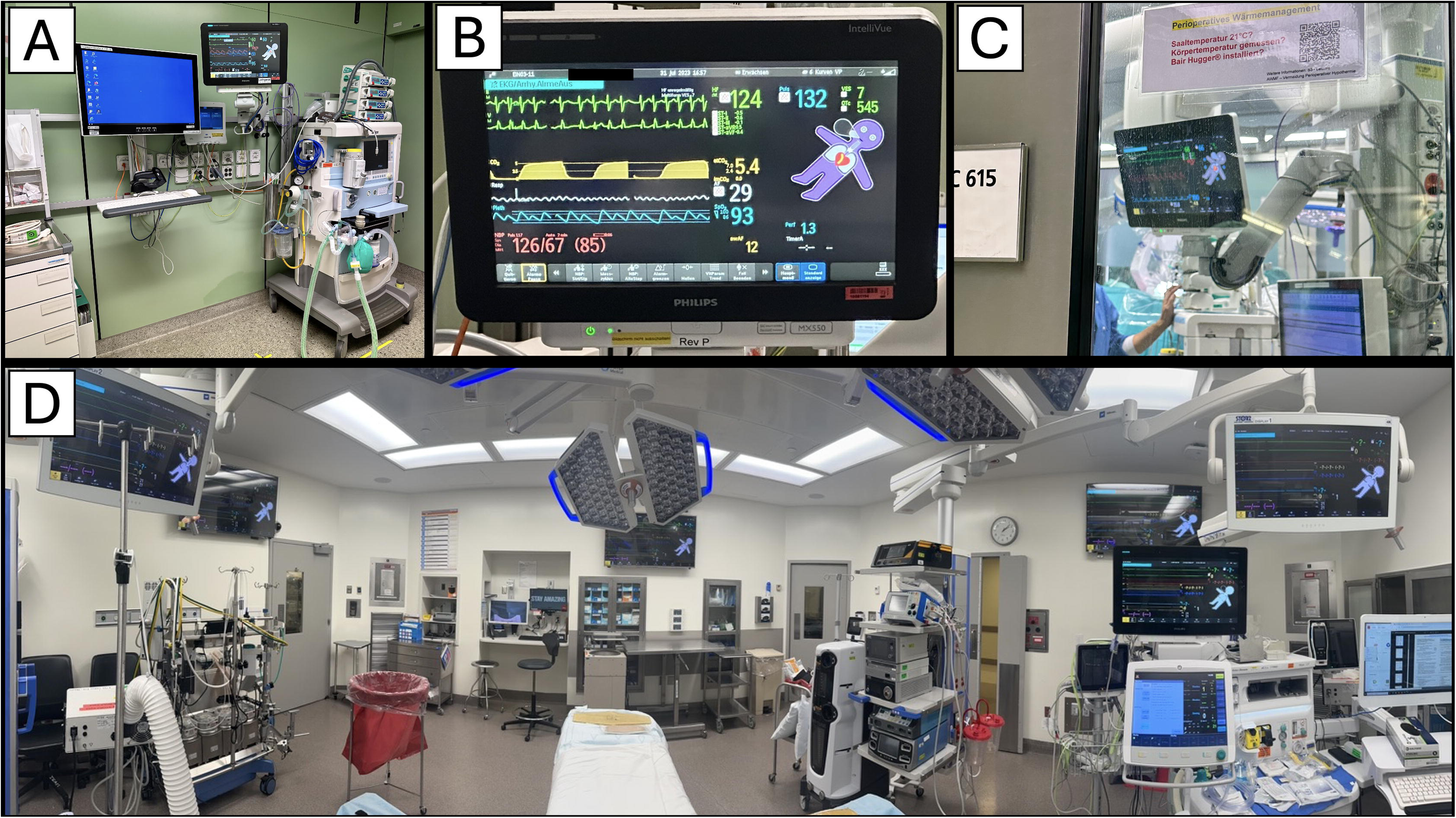
Conventional patient monitoring before and after implementation of the Visual Patient Avatar. **A:** Conventional patient monitoring display prior to implementation of the Visual Patient Avatar (VPA), showing physiological information exclusively as numerical values and waveforms on a Philips IntelliVue monitor. **B**: Identical monitoring configuration after implementation of the VPA, with the avatar-based visualisation displayed alongside conventional numerical values and waveforms. The VPA integrates multiple vital signs and sensor states into a single animated virtual patient to support clinicians’ situation awareness, while preserving the full conventional monitoring display.

**Figure 2:**
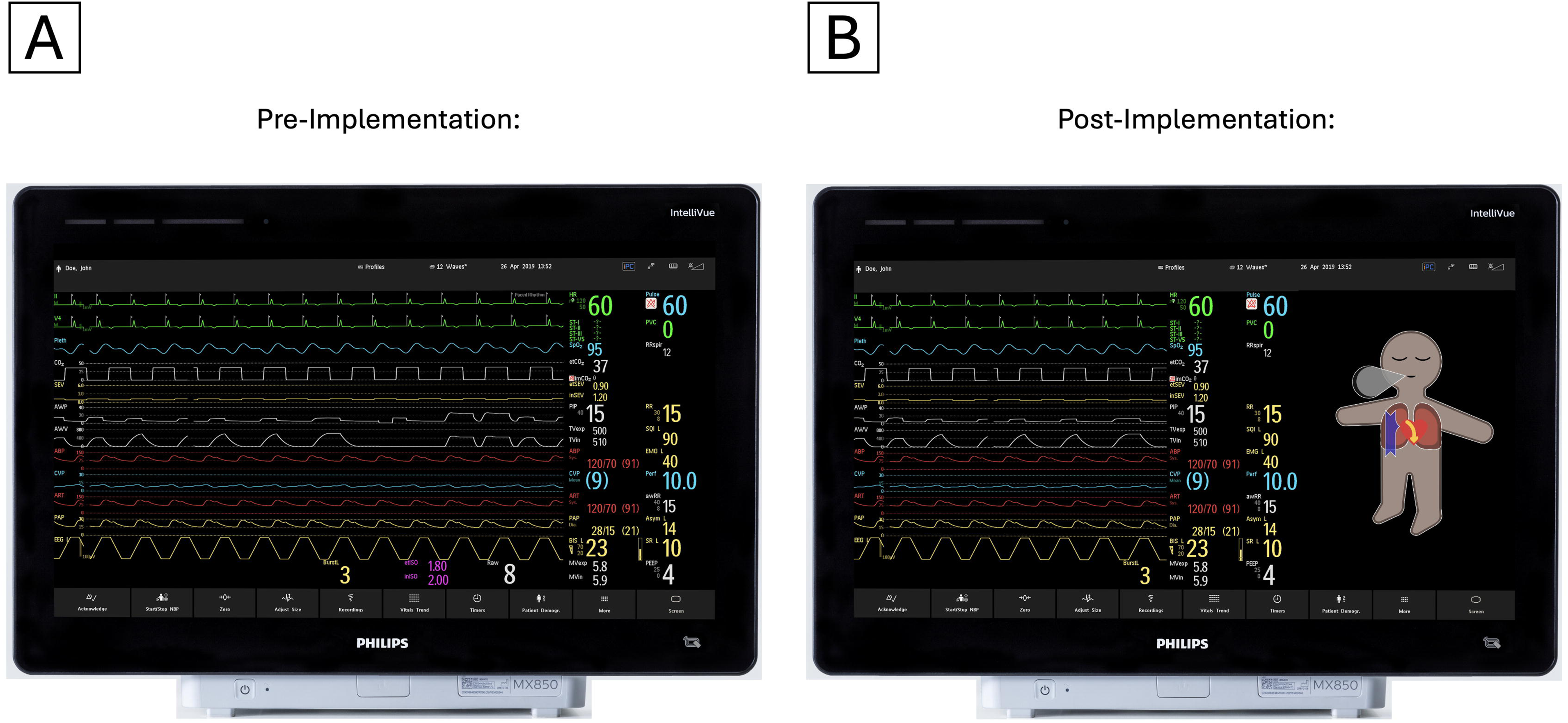
Clinical deployment of the Visual Patient Avatar (VPA) in routine perioperative monitoring. **A:** Standard anaesthesia workstation setup at the University Hospital Zurich. **B**: Example of a patient monitor showing a hypoxaemic patient. The avatar appears purple, indicating oxygen desaturation, and exhibits rapid pulsation corresponding to the elevated heart rate (not visible in still photo). **C:** Example of a patient monitor depicting a hypothermic patient indicated by snow flake symbols, which are visible from a distance, looking through the glass of the OR door. **D:** Standard monitoring setup in a cardiac operating theatre at New York–Presbyterian Hospital, illustrating the integration of the VPA across multiple wall-mounted and bedside displays.

In addition, clinicians could select predefined care profiles tailored to different clinical contexts (e.g., adult vs. paediatric care). Care profile selection adjusted both conventional alarm thresholds and the corresponding Visual Patient Avatar (VPA) thresholds in a single step, enabling rapid adaptation to the clinical context. After initial profile selection, conventional alarm limits remained fully adjustable by users according to clinical needs, while VPA thresholds were fixed within each care profile to represent the centre-specific standard configuration for that clinical context. All care profiles and centre-specific alarm and VPA threshold settings are provided in **Supplementary Material 1**.

### Implementation process and training

Implementation is preceded by joint planning sessions involving clinical leadership (medical, nursing, patient safety and quality management) and Philips representatives to adapt deployment to local workflows.

The Go-Live phase includes structured training in the use of VPA and the underlying concept of situation awareness, consisting of lectures, bedside teaching and walk-in training stations available during the installation period. After Go-Live, local change champions (practising anaesthetists at each centre) remain available for ongoing support, institutional teaching sessions and bedside assistance as needed.

### Study outcomes

#### Primary outcome

- **Vital sign deviation time**: defined as the cumulative duration during which a given vital parameter exceeded predefined upper or lower thresholds. To allow comparability across patients and monitoring durations, deviation time was normalized to 60 minutes of effective measurement time for the respective vital sign, calculated as **60 × (deviation time / measurement time)**. Here, *measurement time* refers to the time during which valid measurements of the respective vital sign were available in the dataset. Thresholds were centre-specific and reflected local configurations. Outcomes were calculated separately for each vital sign displayed in the VPA **(Table 2)**.

#### Secondary outcomes

- **VitalSign AUC**: the area under the curve of vital sign deviations. Each vital sign has an upper limit *U* and a lower limit *L*, defining the admissible interval [*L*,*U*]. For each minute *i*, the recorded value *v*1_ is compared with this interval, and the absolute deviation *d*1_ from the admissible range is calculated. The VitalSign_AUC is defined as **VitalSign AUC =** Σ**(*d***LJ **× 1 minute)**, capturing both magnitude and duration of deviations from the admissible range.
- Alarm count (per 60 minutes): total number of audible alarms aggregated across all vital signs that were being monitored. Table 2 lists the vital signs that could trigger alarms. Alarm thresholds were configurable by users according to local monitoring settings. To account for differences in monitoring duration and availability of vital signs, alarm counts were normalised to 60 minutes of measurement time using the formula: Alarm count = 60 × (sum of per-vital-sign alarm counts) / (sum of per-vital-sign measurement times).
- Alarm time (per 60 minutes): cumulative duration of audible alarms aggregated across all monitored vital signs. This approach reflects clinical practice, where multiple simultaneous alarms are presented independently on the monitor and contribute cumulatively to alarm burden. Alarm time was normalised to 60 minutes of measurement time using the formula: Alarm time = 60 × (sum of per-vital-sign alarm durations) / (sum of per-vital-sign measurement times).
- Alarm reaction time: mean time from (acknowledged) alarm onset until the alarm was either acknowledged by the user or all alarms were paused through user input, as recorded in the alarm logs. If multiple alarms occurred simultaneously, reaction time was calculated separately for each alarm event.

Not all vital signs have corresponding acoustical alarms or sufficient temporal resolution, details are provided in Table 2 footnotes.

**Table 2:** Adult threshold settings used at New York–Presbyterian/Weill Cornell Medicine and University Hospital Zurich. Vital sign thresholds are centre-specific and reflect the respective local alarm configurations used in routine clinical monitoring. At each participating centre, these thresholds are defined and configured by local clinical expert groups according to established clinical practice. The thresholds were used to calculate Vital Sign Deviation Time and VitalSign AUC. In addition, the vital signs listed in Table 2 were used for the calculation of the secondary alarm-related outcomes, including alarm count, alarm time, and alarm reaction time.

| Vital sign | Low thresholds | High thresholds | Units |
| --- | --- | --- | --- |
| **NewYork-Presbyterian/Weill Cornell** |  |  |  |
| Heart rate (ECG) | 45 | 135 | /min |
| Pulse | 45 | 135 | /min |
| Blood Pressure (Mean) | 55 | 125 | mmHg |
| CVP (Mean) | 2 | 14 | mmHg |
| STE | none | none | mV |
| SpO2 | 94 | 100 | % |
| Respiratory rate | 10 | 25 | /min |
| Tidal Volume | 200 | 1200 | ml |
| etCO2 | 25 | 55 | mmHg |
| Body temperature | 35.5 | 38 | deg C |
| BIS | <65 | ≥65 | N/A |
| TOF ratio | <90 | ≥90 | % |
| Cardiac index | 4.5 | 8 | L/min/<br>m<sup>2</sup> |
| Peak airway pressure | 5 | 35 | mbar |
| FiO2 | 21 | 80 | % |
| **University Hospital Zurich** |  |  |  |
| Heart rate (ECG) | 55 | 115 | /min |
| Pulse<sup>a</sup> | 55 | 115 | /min |
| Blood Pressure (Mean) | 65 | 105 | mmHg |
| CVP (Mean) | 4 | 12 | mmHg |
| STE | <-0.2 | >0.2 | mV |
| SpO2 | 94 | none | % |
| Respiratory rate | 8 | 16 | /min |
| Tidal Volume<sup>b</sup> | 250 | 750 | ml |
| etCO2 | 4.2 | 5.5 | kPa |
| Body temperature<sup>c</sup> | 36.1 | 37.5 | °C |
| BIS | <55 | none | (-) |
| TOF ratio<sup>a</sup> | <90 | none | % |
| Peak airway pressure | 10 | 30 | mbar |

### Rationale for the chosen outcome measures

Deviations of vital signs beyond predefined thresholds as well as frequent and prolonged audible alarms are known to compromise patient *outcomes and patient* safety in perioperative care. Both centres have defined institution-specific safe ranges for vital signs and corresponding alarm configurations that reflect local standards of care.

Not all vital signs are associated with audible alarms by default, and alarms may be paused or adapted by users during clinical care, whereas the underlying vital-sign thresholds remain fixed. For this reason, both vital-sign–based outcomes and audible alarm–related outcomes are evaluated to obtain a comprehensive assessment of the effects of VPA.

Because VPA was explicitly designed to improve clinicians’ situation awareness of patients’ vital-sign status, these outcomes are considered well suited to detect potential improvements associated with the intervention.

### Data sources and data collection

At NYP/WCM, monitoring and alarm data are recorded through the institutional data warehouse infrastructure, providing high-resolution (1 Hz) vital sign and alarm data linked to the anaesthesia record. Data extraction and linkage are performed by the NYP/WCM study and data warehouse teams.

At USZ, vital signs are captured via CapsuleTech nodes at a sampling rate of 1 minute, and case-level metadata are obtained from the institutional data lake (IMDSoft MetaVision). Audible alarm logs are downloaded from Philips PICiX central stations at a resolution of 1 Hz. Planned variables and their respective source systems are summarised in **Table 3**.

**Table 3:** Data acquisition per case and the sources of data in Zurich. PDMS: Patient data management system. DWC=Philips Data Warehouse Connect, ASA=American Society of Anesthesiologists, AUC=Area under the curve, PICiX=Philips Central Station.

| **Data** | **Measurement** | **NYP** | **USZ** |
| --- | --- | --- | --- |
| **Demographic data** |  |  |  |
| Study phase | Categorical | DWC | Data lake |
| End of anaesthesia | Time in seconds | DWC | data lake |
| End of surgery | Time in seconds | DWC | data lake |
| Gender (used for adjustment) | Categorical | DWC | data lake |
| Age (used for adjustment) | Categorical | DWC | data lake |
| Body Mass Index (used for adjustment) | Categorical | DWC | data lake |
| ASA classification (used for adjustment) | Categorical | DWC | data lake |
| Surgical risk classification (used for adjustment)<br>A 1–6 scale reflecting the inherent risk of the intervention, with higher values indicating higher complexity and risk. | Categorical | DWC | data lake |
| Urgency of surgery (used for adjustment) | Categorical | DWC | data lake |
| Type of surgery | Categorical | DWC | data lake |
| Surgical specialty | Categorical | DWC | data lake |
| Type of anesthesia (used for adjustment) | Categorical | DWC | data lake |
| Type of airway device | Categorical | DWC | data lake |
| Type of regional anesthesia | Categorical | DWC | data lake |
| **Vital signs data** |  |  |  |
| Vital sign deviation | Time in min | DWC | data lake |
| Vital sign time-based Area Under the Curve (AUC) | dimensionless AUC | DWC | data lake |
| **Alarms data** |  |  |  |
| Alarm count | Numeric | DWC | PICiX |
| Alarms time | Time in seconds | DWC | PICiX |
| Alarm reaction time | Time in seconds | DWC | PICiX |

### Data management, security and confidentiality

Data management is conducted by trained and authorised personnel at each centre in accordance with local policies, data governance requirements and regulatory approvals. Access is restricted to approved study personnel, and all data transfers use secure institutional mechanisms.

### Measures to ensure data quality

Data extraction relies on established routine clinical documentation and monitoring pipelines at both centres. Quality checks are performed during data export and prior to statistical analysis and include completeness checks, plausibility range checks, timestamp consistency and verification of successful data linkage.

### Statistical analysis

The unit of analysis is the individual anaesthesia case. Outcomes are computed at the case level and analyzed separately for each vital sign (and for each alarm-related outcome). The analysis compares the pre-implementation phase versus the post-implementation phase; the sink-in phase is excluded from the analysis.

Case characteristics will be summarized using means (SD) or medians (IQR) for continuous variables and counts (%) for categorical variables. Descriptive comparisons between phases may use t-tests/Wilcoxon tests for continuous variables and chi-square/Fisher’s exact tests for categorical variables.

For each outcome, multivariable regression models will be used to estimate associations between implementation phase (post vs pre) and outcomes, adjusting for prespecified covariates (see Table 3). Outcomes are normalized to 60 minutes of case time. Depending on the outcome, if necessary, data sources will be joined. Alarms without corresponding measurement time are considered an error and will be dropped.

Model choice will be guided by outcome distribution and diagnostics. Linear regression will be used as the default; if residual diagnostics suggest marked non-normality or heteroskedasticity, outcomes may be log-transformed (with a small constant added if needed to accommodate zeros) or modeled using generalized linear models (e.g., Gamma regression with log link for right-skewed positive outcomes). If an outcome exhibits a large point mass at zero, two-part or zero-inflated models may be considered.

To address multiple testing issues, we will apply Bonferroni correction within each study outcome separately (thus, the family-wise error rate for all the tests using the same study outcome is controlled at 5%). Analyses will be performed separately for each centre.

For each outcome, analyses will include cases with non-missing outcome data. Missingness patterns will be described. For covariates, if the proportion of missing values is ≤1%, the observations will be dropped. If >1%, multiple imputation by chained equations (MICE) will be used, and regression estimates will be pooled using Rubin’s rules.

### Sample size considerations

This study leverages routinely collected data from large perioperative case volumes at both centres. Formal power calculations are not performed because the sample size is determined by clinical caseloads during the prespecified phase windows. The anticipated large sample supports precise estimation of associations and enables stratified analyses by care area and centre.

### Ethics and dissemination

#### Ethics and data governance approvals

The study was approved by the Institutional Review Board of Weill Cornell Medicine (IRB No. 22-05024803).

The Cantonal Ethics Committee Zurich issued a declaration of non-jurisdiction in accordance with Swiss regulations (BASEC Req-2021-00756; 6 July 2021). At USZ, the institutional Data Governance Board approved the project, including the data export and anonymisation workflow (DGR0000504; 31 January 2025). A collaboration and data-sharing agreement between Weill Cornell Medicine and the University of Zurich was established on 11 April 2023.

### Data deposition and curation

Data supporting the findings of this study are derived from routinely collected anonymised perioperative monitoring and alarm data recorded at the participating institutions during the prespecified study periods.

Anonymized data and statistical code may be made available from the corresponding author upon reasonable request and subject to approval by the participating institutions and relevant data governance bodies.

### Dissemination

Study findings will be reported in peer-reviewed scientific journals and presented at national and international scientific meetings. A primary joint publication including data from both centres is anticipated. Secondary analyses using centre-specific datasets may be published separately following completion of the primary analysis, in accordance with institutional policies and the collaboration agreement.

## Discussion

This protocol describes a pragmatic, dual-centre evaluation of an avatar-based situation awareness display introduced into routine perioperative care. By focusing on objective monitoring metrics (time outside predefined vital sign ranges and audible alarm burden), the study aims to test whether human-factors benefits demonstrated in controlled studies translate into measurable changes in real-world practice. If favourable associations are observed, subsequent studies could explore downstream patient-centred outcomes.

### Strengths and limitations

This study uses large-scale, routinely collected perioperative monitoring data from two tertiary academic centres, enabling evaluation of the VPA under real-world clinical conditions across diverse patient populations, care areas and institutional contexts. The dual-centre design enhances generalisability and allows both centre-specific and comparative analyses.

The quasi-experimental pre–post design does not allow causal inference. Although multivariable regression models adjust for relevant patient- and procedure-related characteristics, residual confounding by time and unmeasured factors cannot be fully excluded. To address learning and adoption effects associated with the introduction of a new monitoring visualisation, a dedicated sink-in phase was incorporated; nevertheless, temporal confounding remains a potential limitation. We assume independence of the individual anaesthesia cases. This assumption might be violated because the same operating team will treat several patients (perhaps even multiple times). However, this information is not available.

Data collection occurred during different calendar periods and healthcare systems, which introduces the possibility that external factors unrelated to the intervention may influence outcomes, including possible staff turnover and new trainees. At the same time, this heterogeneity strengthens the evaluation of the intervention across varied real-world environments.

Alarm configurations and visualisation thresholds differ between centres and reflect local standards of care. While this limits direct standardisation of absolute alarm frequencies, it allows assessment of the intervention across heterogeneous alarm environments without artificial harmonisation.

Finally, the primary outcomes represent process-level measures of physiological stability and alarm burden rather than direct patient outcomes. However, deviations in vital signs and exposure to audible alarms are well-established indicators of patient safety and workload, providing a clinically meaningful basis for evaluating the potential impact of the intervention and informing future outcome-focused research.

## CRediT Author Statement

**SYJ**: Conceptualization, Methodology, Writing – original draft, Writing – review & editing, Supervision, Project administration, Funding acquisition.

**TRR**: Conceptualization, Methodology, Study preparation, Writing – original draft, Writing – review & editing, Supervision, Project administration.

**VET**: Statistical methodology, Software, Writing – review & editing.

**ME**: Methodology, Study preparation, Writing – review & editing.

**MK**: Statistical methodology, Software, Writing – review & editing.

**GD**: Statistical methodology, Software, Writing – review & editing.

**KC**: Statistical methodology, Software, Writing – review & editing.

**LM**: Statistical methodology, Software, Writing – review & editing.

**MT**: Study preparation, Methodology, Writing – review & editing.

**AM**: Study preparation, Methodology, Writing – review & editing.

**CBN**: Conceptualization, Study preparation, Methodology, Writing – review & editing.

**ZAT**: Conceptualization, Methodology, Writing – original draft, Writing – review & editing, Supervision, Project administration, Funding acquisition.

**DWT**: Conceptualization, Methodology, Writing – original draft, Writing – review & editing, Supervision, Project administration, Funding acquisition.

## Data availability statement

Data used in this study consist of routinely collected perioperative patient monitoring and alarm data from two tertiary academic hospitals and are not publicly available due to data protection regulations and their sensitive nature.

The datasets were anonymised prior to analysis in accordance with institutional and regulatory requirements. Access to the data is restricted and subject to approval by the respective institutional data governance bodies.

Data may be made available from the corresponding author upon reasonable request, subject to approval by the University Hospital Zurich Data Governance Board and the Institutional Review Board of Weill Cornell Medicine, and in compliance with applicable data protection laws and institutional policies.

Statistical code used for the analysis may be made available upon reasonable request.

## Competing Interests Statements

Authors CBN and DWT are named inventors of the Visual Patient Avatar technology. This technology has been licensed by the University of Zurich to Philips. In accordance with University of Zurich regulations, inventors are entitled to an appropriate share of licensing revenues, and CBN and DWT receive royalties related to this technology.

The Zurich-based authors TRR, CBN, and DWT have received honoraria and travel support for lectures from Philips.

## Funding

Authors TRR and DWT have received research funding from Philips through a joint development agreement between Philips Medizin Böblingen GmbH and the University of Zurich for the Visual Patient Avatar (dated 18 December 2018). In addition, TRR and DWT received research grants from Philips Healthcare, Philips North America (Exhibit B – USZ-0007; Exhibit B – USZ-0010).

Authors SYJ, VET, and ZAT received research funding from Philips Healthcare, Philips North America (SRA_233936/WCM-0001).

All funding and remuneration were managed through institutional agreements. This is an investigator-initiated study. Philips had no role in the study design, data collection, data analysis, data interpretation, manuscript preparation, or decisions regarding publication.

## Declaration of Generative AI and AI-assisted Technologies in the Writing Process

Generative AI (ChatGPT model 5.2, OpenAI, San Francisco, CA, USA) was used to enhance the clarity and readability of the manuscript. All AI-assisted revisions were reviewed and approved by the authors to ensure accuracy and completeness.

## Notes

### Author Declarations

The study was approved by the Institutional Review Board of Weill Cornell Medicine (IRB No. 22-05024803). The Cantonal Ethics Committee Zurich issued a declaration of non-jurisdiction in accordance with Swiss regulations (BASEC Req-2021-00756; 6 July 2021). At USZ, the institutional Data Governance Board approved the project, including the data export and anonymisation workflow (DGR0000504; 31 January 2025). A collaboration and data-sharing agreement between Weill Cornell Medicine and the University of Zurich was established on 11 April 2023.

