## Supplementary Material 1 for "Study protocol Effects of Philips Visual Patient Avatar on vital sign deviations and audible alarm burden in perioperative care: a dual-centre, quasi-experimental pre-post big-data study protocol (NewYork-Presbyterian/Weill Cornell and University Hospital Zurich)"

**Table of contents:**

**Page 2: Supplementary Figure S1:** Examples of selectable patient monitor screen layouts incorporating the Visual Patient Avatar (VPA).

**Pages 3-6: Care profiles NewYork-Presbyterian/Weill Cornell**

**Pages 7-9: Care profiles University Hospital Zurich**

**Supplementary Figure S1:** Examples of selectable patient monitor screen layouts incorporating the VPA. Panels **A and B** show example screen layouts used at the NewYork-Presbyterian/Weill Cornell study site, while panels **C and D** illustrate example layouts used at the University Hospital Zurich study site. All layouts display the VPA alongside conventional numerical values and physiological waveforms, while varying the relative size, position, and information density of the avatar and conventional monitoring elements. Users were free to select their preferred screen layout according to individual preferences and clinical context throughout routine care.

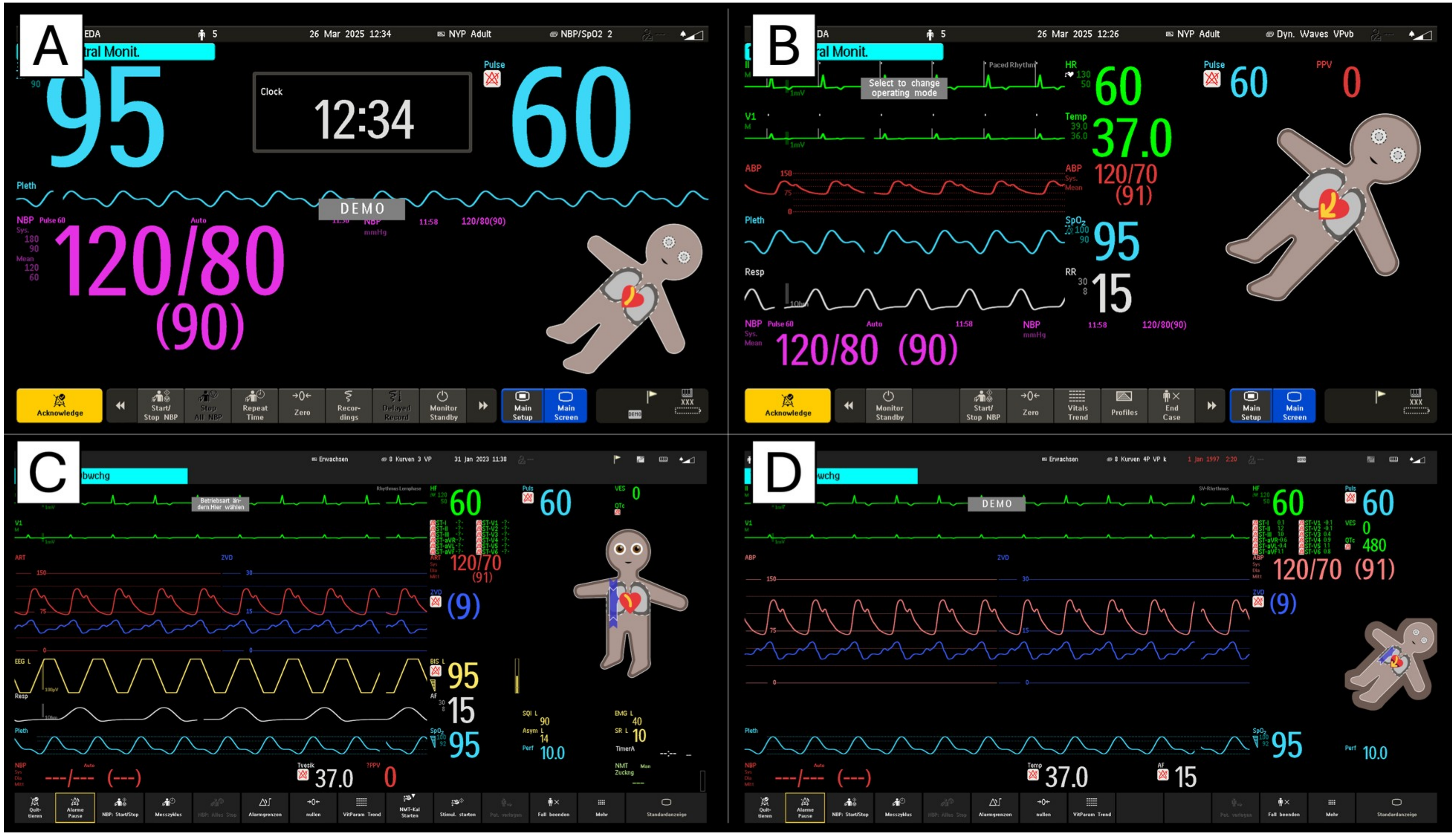

**Care profiles with alarm and Visual Patient Avatar (VPA) thresholds NewYork-Presbyterian/Weill Cornell**

ABP=arterial blood pressure, NBP=non-invasive blood pressure, CVP=central venous pressure, STE=ST-segment elevation, BIS=bispectral index, TOF=Train-of-four, FiO2=Fraction inspiratory oxygen, N/A=not available=none set per default.

| Parameter | Low Alarm | Low VPA | High VPA | High Alarm | Units |
|---|---|---|---|---|---|
| (Adult CVOR) | | | | | |
| ECG(HR) | 40 | 45 | 135 | 140 | bpm |
| Pulse | 40 | 45 | 135 | 140 | bpm |
| ABP (mean) | 50 | 55 | 125 | 130 | mmHg |
| NBP (mean) | 50 | 55 | 125 | 150 | mmHg |
| CVP (Mean) | 0 | 2 | 14 | 16 | mmHg |
| STE | N/A | N/A | N/A | N/A | mV |
| SpO2 | 90 | 94 | 100 | 100 | % |
| Respiratory rate | 8 | 10 | 25 | 30 | bpm |
| Tidal volume | N/A | 200 | 1200 | 1500 | ml |
| etCO2 | 20 | 25 | 55 | 60 | mmHg |
| Body temperature | 35 | 35,5 | 38 | 38,3 | deg C |
| BIS | 20 | N/A | 65 | 70 | N/A |
| TOF ratio | N/A | 90 | 90 | N/A | % |
| Cardiac index | 4 | 4,5 | 8 | 8,5 | l/m^2/min |
| Peak airway pressure | N/A | 5 | 35 | 40 | mbar |
| FiO2 | 20 | 21 | 80 | N/A | % |

| Parameter | Low Alarm | Low VPA | High VPA | High Alarm | Units |
|---|---|---|---|---|---|
| (Adult VAD) | | | | | |
| ECG (HR) | 40 | 45 | 135 | 140 | bpm |
| Pulse | 40 | 45 | 135 | 140 | bpm |
| ABP (mean) | 50 | 55 | 145 | 150 | mmHg |
| NBP (mean) | 50 | 55 | 145 | 150 | mmHg |
| CVP (Mean) | 0 | 2 | 8 | 10 | mmHg |
| STE | N/A | N/A | N/A | N/A | mV |
| SpO2 | 90 | 94 | 100 | 100 | % |
| Respiratory rate | 8 | 10 | 25 | 30 | bpm |
| Tidal volume | 0 | 200 | 1200 | 1500 | ml |
| etCO2 | 20 | 25 | 55 | 60 | mmHg |
| Body temperature | 35 | 35,5 | 38 | 38,3 | deg C |
| BIS | 20 | N/A | 65 | 70 | N/A |
| TOF ratio | N/A | 90 | 90 | N/A | % |
| Cardiac index | 4 | 4,5 | 8 | 8,5 | l/m^2/min |
| Peak airway pressure | N/A | 10 | 35 | 40 | mbar |
| FiO2 | 20 | 21 | 80 | N/A | % |

| Parameter | Low Alarm | Low VPA | High VPA | High Alarm | Units |
|---|---|---|---|---|---|
| (Adult Robot) | | | | | |
| ECG (HR) | 40 | 45 | 135 | 140 | bpm |
| Pulse | 40 | 45 | 135 | 140 | bpm |
| ABP (mean) | 50 | 55 | 125 | 130 | mmHg |
| NBP (mean) | 50 | 55 | 125 | 150 | mmHg |
| CVP (Mean) | 0 | 2 | 14 | 16 | mmHg |
| STE | N/A | N/A | N/A | N/A | mV |
| SpO2 | 90 | 94 | 100 | 100 | % |
| Respiratory rate | 8 | 10 | 25 | 30 | bpm |
| Tidal volume | N/A | 200 | 1200 | 1500 | ml |
| etCO2 | 20 | 25 | 55 | 60 | mmHg |
| Body temperature | 35 | 35,5 | 38 | 38,3 | deg C |
| BIS | 20 | N/A | 65 | 70 | N/A |
| TOF ratio | N/A | 90 | 90 | N/A | % |
| Cardiac index | 4 | 4,5 | 8 | 8,5 | l/m^2/min |
| Peak airway pressure | N/A | 5 | 35 | 40 | mbar |
| FiO2 | 20 | 21 | 80 | N/A | % |

| Parameter | Low Alarm | Low VPA | High VPA | High Alarm | Units |
|---|---|---|---|---|---|
| (Adult Bypass) | | | | | |
| ECG (HR) | 40 | 45 | 135 | 140 | bpm |
| Pulse | 40 | 45 | 135 | 140 | bpm |
| ABP (mean) | 50 | 55 | 145 | 150 | mmHg |
| NBP (mean) | 50 | 55 | 145 | 150 | mmHg |
| CVP (Mean) | -2 | 0 | 24 | 26 | mmHg |
| STE | N/A | N/A | N/A | N/A | mV |
| SpO2 | 90 | 94 | 100 | 100 | % |
| Respiratory rate | 8 | 10 | 25 | 30 | bpm |
| Tidal volume | N/A | 200 | 1200 | 1500 | ml |
| etCO2 | 20 | 25 | 55 | 60 | mmHg |
| Body temperature | 0 | 35,5 | 38 | 38,3 | deg C |
| BIS | 20 | N/A | 65 | 70 | N/A |
| TOF ratio | N/A | 90 | 90 | N/A | % |
| Cardiac index | 4 | 4,5 | 8 | 8,5 | l/m^2/min |
| Peak airway pressure | N/A | 5 | 35 | 40 | mbar |
| FiO2 | 20 | 21 | 80 | N/A | % |

| Parameter | Low Alarm | Low VPA | High VPA | High Alarm | Units |
|---|---|---|---|---|---|
| **Adult NBP SpO2** | | | | | |
| **ECG (HR)** | 50 | 55 | 115 | 120 | bpm |
| **Pulse** | 50 | 55 | 115 | 120 | bpm |
| **ABP (mean)** | 70 | 65 | 105 | 110 | mmHg |
| **NBP (mean)** | 60 | 65 | 105 | 120 | mmHg |
| **CVP (Mean)** | 0 | 4 | 12 | 10 | mmHg |
| **STE** | N/A | -0,2 | 0,2 | N/A | mV |
| **SpO2** | 90 | 94 | N/A | 100 | % |
| **Respiratory rate** | 8 | 10 | 16 | 30 | bpm |
| **Tidal volume** | N/A | 250 | 750 | N/A | ml |
| **etCO2** | 25 | N/A | N/A | 60 | mmHg |
| **Body temperature** | 35 | 36,1 | 37,5 | 39 | deg C |
| **BIS** | 20 | 55 | 55 | 70 | N/A |
| **TOF ratio** | N/A | 90 | 90 | N/A | % |
| **Cardiac index** | N/A | 2,5 | 4 | N/A | l/m^2/min |
| **Peak airway pressure** | N/A | 10 | 30 | N/A | mbar |
| **FiO2** | N/A | 80 | 80 | N/A | % |

| Parameter | Low Alarm | Low VPA | High VPA | High Alarm | Units |
|---|---|---|---|---|---|
| **Neo** | | | | | |
| **ECG (HR)** | 100 | 105 | 195 | 200 | bpm |
| **Pulse** | 100 | 105 | 195 | 200 | bpm |
| **ABP (mean)** | 55 | 60 | 85 | 90 | mmHg |
| **NBP (mean)** | 40 | 60 | 85 | 90 | mmHg |
| **CVP (Mean)** | 0 | 1 | 3 | 4 | mmHg |
| **STE** | N/A | N/A | N/A | N/A | mV |
| **SpO2** | 85 | 87 | 93 | 95 | % |
| **Respiratory rate** | 30 | 35 | 95 | 100 | rpm |
| **Tidal volume** | N/A | 10 | 40 | 1500 | ml |
| **etCO2** | 20 | 25 | 55 | 60 | mmHg |
| **Body temperature** | 35,5 | 36 | 37,5 | 38,3 | deg C |
| **BIS** | 20 | N/A | 65 | 70 | N/A |
| **TOF ratio** | N/A | N/A | 90 | N/A | % |
| **Cardiac index** | 0,3 | 0,5 | 1,1 | 1,3 | l/m^2/min |
| **Peak airway pressure** | N/A | 1 | 25 | 40 | mbar |
| **FiO2** | 20 | 23 | 100 | N/A | % |

| Parameter | Low Alarm | Low VPA | High VPA | High Alarm | Units |
|---|---|---|---|---|---|
| **Pedi** | | | | | |
| **ECG (HR)** | 90 | 95 | 145 | 150 | bpm |
| **Pulse** | 90 | 95 | 145 | 150 | bpm |
| **ABP (mean)** | 80 | 85 | 125 | 130 | mmHg |
| **NBP (mean)** | 80 | 85 | 125 | 130 | mmHg |
| **CVP (Mean)** | 0 | 2 | 8 | 10 | mmHg |
| **STE** | N/A | N/A | N/A | N/A | mV |
| **SpO2** | 95 | 95 | 95 | 100 | % |
| **Respiratory rate** | 20 | 25 | 55 | 60 | rpm |
| **Tidal volume** | N/A | 20 | 400 | 1500 | ml |
| **etCO2** | 20 | 25 | 55 | 60 | mmHg |
| **Body temperature** | 35,5 | 35,7 | 37,5 | 38,3 | deg C |
| **BIS** | 20 | N/A | 65 | 70 | N/A |
| **TOF ratio** | N/A | 90 | 90 | N/A | % |
| **Cardiac index** | 2,6 | 2,8 | 3,5 | 3,7 | l/m^2/min |
| **Peak airway pressure** | N/A | 1 | 27 | 40 | mbar |
| **FiO2** | 20 | 23 | 100 | N/A | % |

**Care profiles with alarm and VPA thresholds University Hospital Zurich**

ABP=arterial blood pressure, NBP=non-invasive blood pressure, CVP=central venous pressure, STE=ST-segment elevation, BIS=bispectral index, TOF=Train-of-four, FiO2=Fraction inspiratory oxygen. N/A=not available=none set per default.

| Parameter | Low alarm | Low VPA | High VPA | High alarm | (Units) |
|---|---|---|---|---|---|
| **(Adult)** | | | | | |
| **Heart rate (ECG)** | 50 | 55 | 115 | 120 | /min |
| **Pulse** | 50 | 55 | 115 | 120 | /min |
| **ABP (Mean)** | 60 | 65 | 105 | 110 | mmHg |
| **NBP (Mean)** | 60 | 65 | 105 | 110 | mmHg |
| **CVP (Mean)** | N/A | 4 | 12 | N/A | mmHg |
| **STE** | N/A | STE thresholds | STE thresholds | N/A | mV |
| **SpO2** | 92 | 94 | N/A | N/A | % |
| **Respiratory rate** | 6 | 8 | 16 | 18 | /min |
| **Tidal volume** | N/A | 250 | 750 | N/A | ml |
| **etCO2** | N/A | 4.2 | 5.5 | N/A | kPa |
| **Body temperature** | N/A | 36.1 | 37.5 | N/A | °C |
| **BIS** | N/A | 55 | 55 | N/A | (-) |
| **TOF ratio.** | N/A | 90 | 90 | N/A | % |
| **Cardiac index** | N/A | 2.5 | 4 | N/A | l/m^2/ min |
| **Airway pressure (Peak)** | N/A | 10 | 30 | N/A | mbar |
| **FiO2** | N/A | 80 | 80 | N/A | % |

**VPA thresholds of ST-Segments:**

| | |
|---|---|
| STE Female V1, V4-6 | 0.8mm |
| STE Female V2, V3 | 1.3mm |
| STE Female V7-V9 | 0.3mm |
| STE Female V3R-V6R | 0.3mm |
| STE Female Limb | 0.8mm |
| STE Male V1, V4-6 | 0.8mm |
| STE Male V2, V3 | 1.8mm |
| STE Male V7-V9 | 0.3mm |
| STE Male V3R-V6R | 0.3mm |
| STE Male Limb | 0.8mm |

| Parameter | Low alarm | Low VPA | High VPA | High alarm | (Units) |
|---|---|---|---|---|---|
| **0-1 month (Neo)** | | | | | |
| **Heart rate (ECG)** | 100 | 105 | 195 | 200 | /min |
| **Pulse** | 100 | 105 | 195 | 200 | /min |
| **ABP (Mean)** | 34 (mean) | 40 | 65 | 70 (mean) | mmHg |
| **NBP (Mean)** | 50 (syst) | 40 | 65 | 90 (syst) | mmHg |
| **NBP (Syst = SBP)** | 50 (syst) | 60 | 85 | 90 (syst) | mmHg |
| **CVP (Mean)** | N/A | 1 | 7 | N/A | mmHg |
| **STE** | N/A | N/A | N/A | N/A | mV |
| **SpO2** | 86 | 87 | 93 | 95 | % |
| **Respiratory rate (only PACU)** | 30 | 35 | 80 | 85 | /min |
| **Tidal volume** | N/A | 20 | 40 | N/A | ml |
| **etCO2** | N/A | 4,5 | 6,0 | N/A | kPa |
| **Body temperature** | N/A | 36,5 | 38,5 | N/A | °C |
| **BIS** | N/A | 55 | 55 | N/A | (-) |
| **TOF ratio.** | N/A | 90 | 90 | N/A | % |
| **Cardiac index** | N/A | | | | l/m^2/min |
| **Airway pressure (Peak)** | N/A | 4 | 20 | N/A | mbar |
| **FiO2** | N/A | 30 | 80 | N/A | % |

| Parameter | Low alarm | Low VPA | High VPA | High alarm | (Units) |
|---|---|---|---|---|---|
| **1-12 month (Infant)** | | | | | |
| **Heart rate (ECG)** | 90 | 95 | 175 | 180 | /min |
| **Pulse** | 90 | 95 | 175 | 180 | /min |
| **ABP (Mean)** | 44 (mean) | 50 | 70 | 75 (mean) | mmHg |
| **NBP (Mean)** | 60 (syst) | 50 | 70 | 110 (syst) | mmHg |
| **NBP (Syst = SBP)** | 60 (syst) | 70 | 105 | 110 (syst) | mmHg |
| **CVP (Mean)** | N/A | 2 | 10 | N/A | mmHg |
| **STE** | N/A | N/A | N/A | N/A | mV |
| **SpO2** | 92 | 92 | 100 | N/A | % |
| **Respiratory rate (only PACU)** | 20 | 25 | 60 | 65 | /min |
| **Tidal volume** | N/A | 30 | 75 | N/A | ml |
| **etCO2** | N/A | 4,2 | 6,0 | N/A | kPa |
| **Body temperature** | N/A | 36,5 | 38,5 | N/A | °C |
| **BIS** | N/A | 55 | 55 | N/A | (-) |
| **TOF ratio.** | N/A | 90 | 90 | N/A | % |
| **Cardiac index** | N/A | | | | l/m^2/min |
| **Airway pressure (Peak)** | N/A | 6 | 20 | N/A | mbar |
| **FiO2** | N/A | 30 | 80 | N/A | % |

| Parameter | Low alarm | Low VPA | High VPA | High alarm | (Units) |
|---|---|---|---|---|---|
| **1-6 years (Preschool)** | | | | | |
| **Heart rate (ECG)** | 70 | 75 | 115 | 120 | /min |
| **Pulse** | 70 | 75 | 115 | 120 | /min |
| **ABP (Mean)** | 44 (mean) | 50 | 75 | 80 (mean) | mmHg |
| **NBP (Mean)** | 70 (syst) | 50 | 75 | 120 (syst) | mmHg |
| **NBP (Syst = SBP)** | 70 (syst) | 80 | 110 | 120 (syst) | mmHg |
| **CVP (Mean)** | N/A | 2 | 12 | N/A | mmHg |
| **STE** | N/A | N/A | N/A | N/A | mV |
| **SpO2** | 92 | 92 | 100 | N/A | % |
| **Respiratory rate (only PACU)** | 15 | 20 | 35 | 40 | /min |
| **Tidal volume** | N/A | 55 | 150 | N/A | ml |
| **etCO2** | N/A | 4,2 | 6,0 | N/A | kPa |
| **Body temperature** | N/A | 36,5 | 38,5 | N/A | °C |
| **BIS** | N/A | 55 | 55 | N/A | (-) |
| **TOF ratio.** | N/A | 90 | 90 | N/A | % |
| **Cardiac index** | N/A | | | | l/m^2/min |
| **Airway pressure (Peak)** | N/A | 8 | 25 | N/A | mbar |
| **FiO2** | N/A | 30 | 80 | N/A | % |

| Parameter | Low alarm | Low VPA | High VPA | High alarm | (Units) |
|---|---|---|---|---|---|
| **6-12 years (School)** | | | | | |
| **Heart rate (ECG)** | 65 | 70 | 115 | 120 | /min |
| **Pulse** | 65 | 70 | 115 | 120 | /min |
| **ABP (Mean)** | 50 (mean) | 55 | 85 | 90 (mean) | mmHg |
| **NBP (Mean)** | 80 (syst) | 55 | 85 | 130 (syst) | mmHg |
| **NBP (Syst = SBP)** | 80 (syst) | 85 | 120 | 130 (syst) | mmHg |
| **CVP (Mean)** | N/A | 4 | 12 | N/A | mmHg |
| **STE** | N/A | N/A | N/A | | mV |
| **SpO2** | 92 | 92 | 100 | N/A | % |
| **Respiratory rate (only PACU)** | 10 | 15 | 30 | 35 | /min |
| **Tidal volume** | N/A | 100 | 300 | N/A | ml |
| **etCO2** | N/A | 4,2 | 6,0 | N/A | kPa |
| **Body temperature** | N/A | 36,2 | 38,5 | N/A | °C |
| **BIS** | N/A | 55 | 55 | N/A | (-) |
| **TOF ratio.** | N/A | 90 | 90 | N/A | % |
| **Cardiac index** | N/A | | | | l/m^2/min |
| **Airway pressure (Peak)** | N/A | 8 | 25 | N/A | mbar |
| **FiO2** | N/A | 30 | 80 | N/A | % |
